## Supporting Information for "Protect or prevent? A practicable framework for the dilemmas of COVID-19 vaccine prioritization"

March 2023

**1** Department of Electrical and Systems Engineering, University of Pennsylvania, Philadelphia, PA, United States

**2** Division of Infectious Diseases, Department of Medicine, University of Pennsylvania School of Medicine, Philadelphia, PA, United States

***** Corresponding Author (rarghal@seas.upenn@edu)

### 1 Symbol Tables and Values

The table below summarizes relevant system notation.

Table 1: System Notation

| Symbol | Description |
|---|---|
| $N$ | Total number of individuals |
| $S_i(t)$ | Fraction of group $i$ susceptible individuals at time $t$ |
| $E_i(t)$ | Fraction of group $i$ exposed individuals at time $t$ |
| $P_i(t)$ | Fraction of group $i$ pre-symptomatic individuals at time $t$ |
| $A_i(t)$ | Fraction of group $i$ asymptomatic individuals at time $t$ |
| $I_i(t)$ | Fraction of group $i$ early-stage infected individuals at time $t$ |
| $L_i(t)$ | Fraction of group $i$ late-stage infected individuals at time $t$ |
| $H_i(t)$ | Fraction of group $i$ hospitalized individuals at time $t$ |
| $V_i(t)$ | Fraction of group $i$ vaccinated individuals at time $t$ |
| $R_i(t)$ | Fraction of group $i$ recovered individuals at time $t$ |
| $D_i(t)$ | Fraction of group $i$ deceased individuals at time $t$ |
| $V_0$ | Vaccination capacity constraint |

Here we present the value(s) of the disease parameters used throughout our investigation (unless a change is otherwise specified) along with the relevant sources. Note that we also analyzed the robustness of our optimal policies to noisy estimates of such parameters in Section 4.3. Parameters governing vaccination properties can be found in Table 3.

Table 2: Disease Parameters

| | Description | Value(s) | Ref. |
|---|---|---|---|
| $\tau$ | Rate of transition out of exposed phase | 4 | [1] |
| $s_i$ | Probability of becoming pre-symptomatic after exposure for X,Y,Z | {0.4,0.8,0.4} | [2],[3] |
| $\eta$ | Rate of transition out of pre-symptomatic phase | 2 | [4] |
| $\rho$ | Transmissibility of pre-symptomatic individuals | 0.8 | [5] |
| $\psi$ | Rate of transition out of asymptomatic phase | 10 | [6] |
| $\mu$ | Transmissibility of asymptomatic individuals | 0.1 | [5] |
| $\phi$ | Rate of transition out of early-stage infected phase | 3 | [4] |
| $\omega$ | Transmissibility of infected individuals | 0.7 | [5] |
| $\zeta$ | Rate of transition out of late-stage infected phase | 3 | [2] |
| $\pi_i$ | Probability of hospitalization after late-stage infection for X,Y,Z | {0.19,0.57,0.19} | [7] |
| $\sigma$ | Rate of transition out of hospitalized phase | 11 | [7],[3] |
| $\lambda_i$ | Probability of death after hospitalization for X,Y,Z | {0.027,0.3,0.027} | [8],[3] |

### 2 Generalizations of the Basic Model

For simplicity of exposition, in the main body we presented a bare bones model in which 1) vaccinated or recovered individuals are not infected, and 2) vaccine has only one dose. In truth, vaccinated and recovered individuals can be infected, albeit with lower rates of infection, symptomatic infection, hospitalization, and death [9, 10]. Also, while some vaccines have one dose, others need two doses [11]. We generalize our model to consider 1) infections after vaccination and recovery (Section 2.1) 2) multi dose vaccines (Section 2.2) and 3) several public health objectives of interest other than overall death count (Section 2.3). The ease of these generalizations demonstrate the flexibility of our model. These generalizations were used throughout our numerical investigation to compute and evaluate the optimal and near-optimal vaccination strategies in Section 4. Similarly, one may allow for vaccination of exposed or infected individuals, ongoing booster vaccines, or other circumstances.

#### 2.1 Breakthrough Infections and Reinfection

We now consider that vaccinated individuals can be infected, though with lower rates of infection, symptomatic infection, hospitalization, and death [9, 10]. Thus, the disease spread rate is lower when at least one of the individuals in the pair involved in transmitting the virus is vaccinated than when neither is vaccinated, transition rates to asymptomatic states are higher for the vaccinated and transition rates to pre-symptomatic, hospitalized, dead are lower for the vaccinated. To allow for the differences in these rates for the vaccinated individuals, a new set of exposed, pre-symptomatic, asymptomatic, early stage infection, late stage infection, hospitalized are created for the vaccinated (Figure S1). The ODE representation of the state dynamics is similar to Figure 1, with the difference that there are now additional states, additional differential equations and additional quadratic terms (i.e. additional red terms) in the differential equations representing disease transmission to the vaccinated individuals. Overall, this leads to 48 differential equations and 48 variables. The optimal control formulation remains the same except that the state trajectories are now provided by the new system of ODEs.

Figure S1 shows the state diagram augmented to allow for breakthrough infection i.e. vaccinated individuals becoming infected. This is done by duplicating the disease progression states to allow for the decreased rates of infection, symptoms, hospitalization, and death in vaccinated individuals (see Table 3).

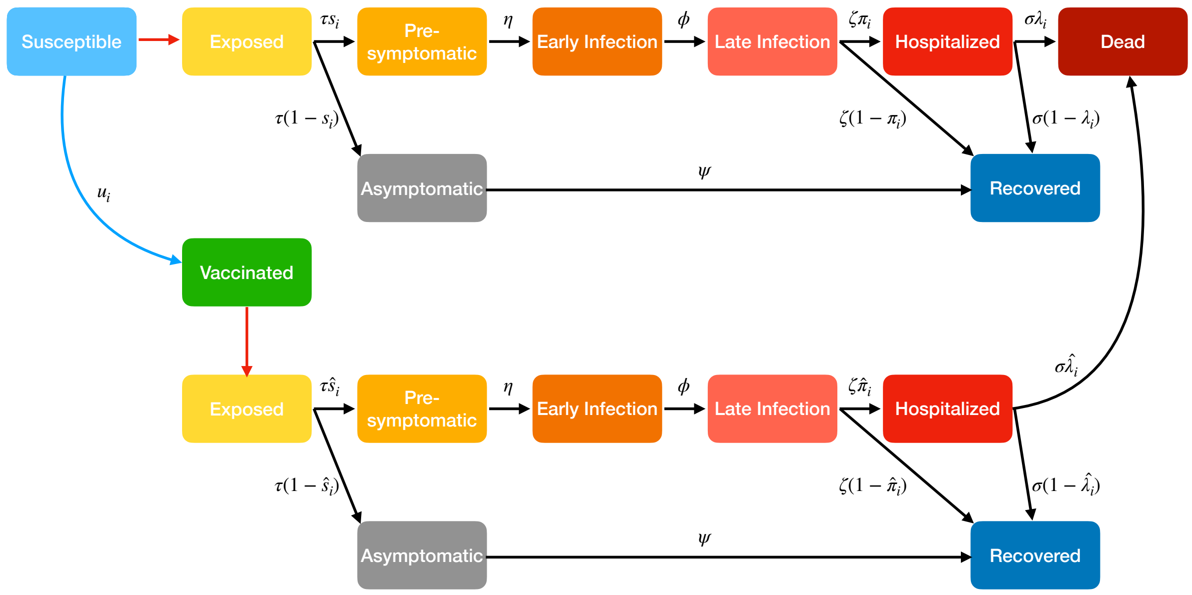


Figure S1: The above diagram depicts the generalized disease states in the single dose vaccine model when expanded to allow for breakthrough infections.

Recovery also provides some degree of immunity from COVID-19, though like vaccination the immunity is not foolproof [9]. To allow for reinfection (i.e. recovered individuals becoming infected), we consider that recovered individuals may become exposed, symptomatic, hospitalized, and deceased at decreased rates by making another copy of the disease states. The set of additional states can model differences in the level of protection afforded via vaccination versus that of acquired immunity.

Both breakthrough infections and reinfection introduce multiplicative factors that represent the decreased rate of exposure, symptoms, hospitalization, and death for vaccinated and recovered individuals [12, 9]. Note that the factors need not be the same for vaccinated and recovered. We now provide the notation and typical values for the respective decreases in the table below along with the relevant sources.

This generalization was used throughout all numerical results presented in Section 4.

Table 3: Vaccine Parameters

| | Description | Value(s) | Ref. |
|---|---|---|---|
| $v_e$ | Multiplicative factor in exposure rate due to vaccination | 0.5 | [12],[13] |
| $v_s$ | Multiplicative factor in symptomatic rate due to vaccination | 0.3 | [12],[13] |
| $v_h$ | Multiplicative factor in hospitalization rate due to vaccination | 0.2 | [12],[13] |
| $v_d$ | Multiplicative factor in death rate due to vaccination | 0.1 | [12],[13] |
| $r_e$ | Multiplicative factor in exposure rate due to reinfection | 0.5 | [9] |
| $r_s$ | Multiplicative factor in symptomatic rate due to reinfection | 0.3 | [9] |
| $r_h$ | Multiplicative factor in hospitalization rate due to reinfection | 0.2 | [9] |
| $r_d$ | Multiplicative factor in death rate due to reinfection | 0.1 | [9] |

#### 2.2 Multi-Dose Vaccination

We also generalize our model to allow for two dose vaccinations. Results for our multi-dose model can be found in Section 4.6. For simplicity we illustrate for the case that an individual does not contract the disease either after recovery or after receiving both doses. These assumptions can be relaxed by expanding the state space as in Section 2.1.

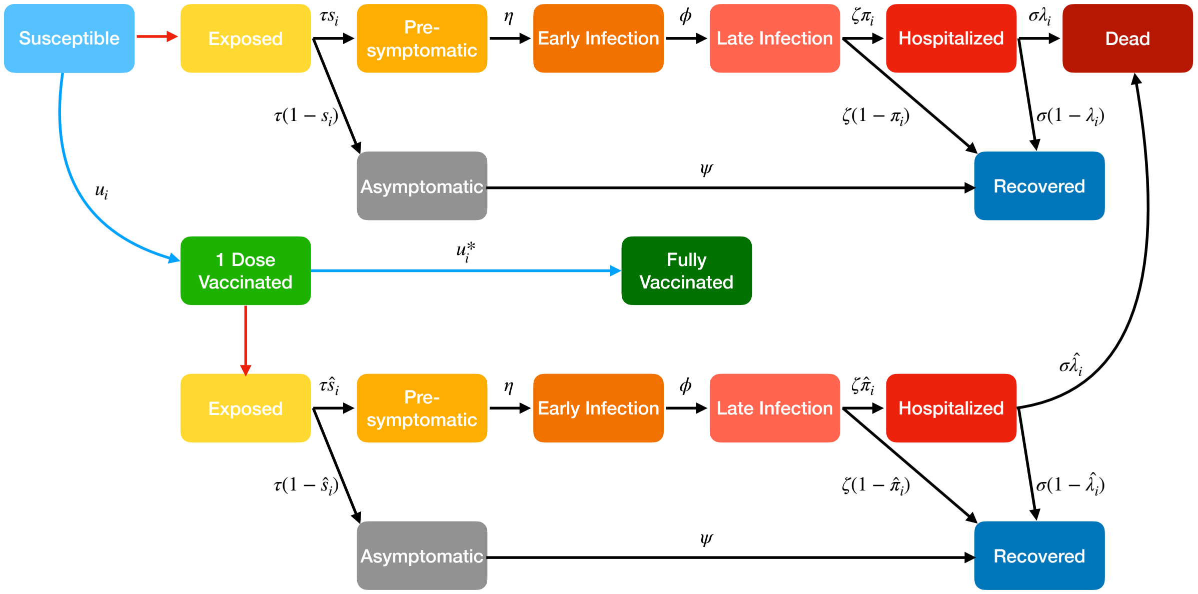


Figure S2: In this expanded two dose state diagram, we introduce the interim "1 Dose Vaccinated" state in which infection is possible but with lower rates as well as decreased risk of symptoms, hospitalization, and death (denoted by $\hat{s}_i < s_i$, $\hat{\pi}_i < \pi_i$, and $\hat{\lambda}_i < \lambda_i$, respectively). Again, for ease of depiction, we present the simplified case in which fully vaccinated and recovered individuals cannot become infected.

We now describe how the ODE formulation for one dose vaccines can be adapted to represent the dynamics when vaccines have two doses. We append to our model an intermediate vaccination state as in Figure S2 in which individuals can be infected, develop symptoms, are hospitalized, and die with lower probability than susceptibles [14, 12, 13]. Susceptibles transition to this state after receiving one dose. After individuals receive both doses they transition from this state to the fully vaccinated state. Let $u_i^*(t)$ refer to the vaccination rate for the second dose of group $i$. This is reflected in similar augmentation to the system of ODEs in Figure 2. We refer to the set of state trajectories for this appended 2 dose system as $\mathcal{S}'$.

Obtaining the optimal vaccination strategy requires the solution of an additional decision problem: the order of the second dose among different groups. For example, should the first dose be administered to as many people as possible first and administering the second dose ought to start only after everyone had received the first dose? Or should an individual be administered the second dose right after the minimum mandated period from the first dose, while others await their first dose? There can be several combinations of the above extremes, eg, depending on which group they belong to, some individuals ought to wait for their second dose until everyone receives the first dose, others receive their second dose as soon as possible after the first dose. We describe the optimal control formulation that will determine the optimal rates for administering first and second doses for different groups and times subject to the capacity constraint for vaccine delivery.

$$\begin{aligned} \text{minimize} \quad & \sum_{i\in\{X,Y,Z\}} D_i(T) \\ \text{subject to} \quad & x \in \mathcal{S}' \\ & x(0) = x_0, \\ & 0 \le u_i(t), u_i^*(t) \le 1 \quad \forall i \in \{X,Y,Z\}, t \in [0,T] \\ & U^*(t) \le V_0 \quad \forall t \in [0,T] \end{aligned} \tag{1}$$

where $U^*(t) = \sum_{i\in\{X,Y,Z\}} S_i u_i + V_i u_i^*$ is the fraction of individuals vaccinated at a given time.

The optimal vaccination strategy can be obtained by solving the above optimal control formulations using the numerical tools described in Section 3.

In 2 dose vaccination, **we define parameter $\alpha$ that denotes the partial protection (as fraction of two dose protection levels in Table 3) afforded by one dose of the vaccine**. That is, the multiplicative factor on exposure after 1 dose will be $1-\alpha(1-v_e)$. We vary $\alpha$ between 0.4 and 0.8 in line with clinical estimates [15]. Findings from our numerical computations using this model are included in Section 4.6.

#### 2.3 Different Objective Functions

Our computational framework is also flexible enough to cater to different public health objectives as shown in Section 4.5. Public health objectives other than the total death count can be minimized by appropriately replacing the objective function. One can minimize the time average of the hospitalization count, total years of life lost (YLL), time average of the symptomatic count, or some combination of these or other metrics of interest. This is noteworthy as an increasing body of work seeks to grapple with the socioeconomic costs of pandemics or even the impacts on social justice [16, 17, 18]. To minimize the time average of the hospitalization count, the objective function in the optimal control formulation in 1 needs to be $\frac{\sum_{i\in\{X,Y,Z\}} \int_0^T H_i(t)dt}{T}$. From the ODEs in Figure 2, $D_i(T) = \sigma\lambda_i \int_0^T H_i(t)dt$. The objective function can therefore be expressed as a weighted sum of the cumulative death count in each group: $\sum_{i\in\{X,Y,Z\}} \frac{D_i(T)}{T\sigma\lambda_i}$. YLLs are computed as a weighted sum of death counts over the different groups where the count in each group is weighted according to the difference between overall life expectancy and the group's mean age (the weight is set to 0 if the difference turns out to be negative). To minimize the time average of the symptomatic count, the objective function has to be $\frac{\sum_{i\in\{X,Y,Z\}} \int_0^T (I_i(t)+L_i(t))dt}{T}$. The time average of the hospitalization counts and symptomatic counts represent number of hospitalized and symptomatic individuals per day respectively; these also respectively represent the number of man days lost due to hospitalization and symptoms normalized by the size of the interval under consideration. The framework as a whole and the optimal control formulation remains the same otherwise.

### 3 Theorem and Proof

We now restate and prove the theorem referred to in Sections 3 and 4.2. For a policy $u(t)$, we will use $U(t)$ to denote the vaccine allocation in absolute quantity rather than fraction i.e. $U_i(t) = u_i(t)S_i(t)$.

**Theorem 1.** *There exists an optimal vaccination strategy $u$ such that, for each $t$ there exist $a(t), b(t), c(t) \in \{X, Y, Z\}$ all distinct and $U_{a(t)}(t) = min\left(V_0, S_{a(t)}(t)\right)$, $U_{b(t)}(t) = min\left(S_{b(t)}(t), V_0 - U_{a(t)}(t)\right)$, and $U_{c(t)}(t) = min\left(S_{c(t)}(t), V_0 - U_{a(t)}(t) - U_{b(t)}(t)\right)$.*

Here we present a proof of Theorem 4.1 in the simplified system which does not allow for reinfection or breakthrough infection. After formulating vaccine prioritization as an optimal control problem, we use Pontryagin's Maximum Principle to obtain necessary conditions for an optimal control from which we discern structural properties [19], [20].

We first formulate the Hamiltonian and Lagrangian as follows:

$$\mathcal{H} := \sum_{i \in \{X,Y,Z\}} (\lambda_i^S \dot{S}_i + \lambda_i^E \dot{E}_i + \lambda_i^P \dot{P}_i + \lambda_i^a \dot{A}_i + \lambda_i^I \dot{I}_i + \lambda_i^L \dot{L}_i + \lambda_i^H \dot{H}_i) \tag{2}$$

$$\mathcal{L} := \mathcal{H} - \mu \left( \sum_{i \in \{X,Y,Z\}} S_i u_i - V_0 \right) \tag{3}$$

where the $\lambda$ costate functions are absolutely continuous and satisfy

$$\dot{\lambda}_i^S = -\frac{\partial \mathcal{L}}{\partial S_i} \quad \dot{\lambda}_i^E = -\frac{\partial \mathcal{L}}{\partial E_i} \quad \dot{\lambda}_i^P = -\frac{\partial \mathcal{L}}{\partial P_i} \quad \dot{\lambda}_i^A = -\frac{\partial \mathcal{L}}{\partial A_i} \quad \dot{\lambda}_i^I = -\frac{\partial \mathcal{L}}{\partial I_i} \quad \dot{\lambda}_i^L = -\frac{\partial \mathcal{L}}{\partial L_i} \quad \dot{\lambda}_i^H = -\frac{\partial \mathcal{L}}{\partial H_i} \tag{4}$$

$$0 = \lambda_i^S(T) \quad 0 = \lambda_i^E(T) \quad 0 = \lambda_i^P(T) \quad 0 = \lambda_i^A(T) \quad 0 = \lambda_i^I(T) \quad 0 = \lambda_i^L(T) \quad 0 = \lambda_i^H(T) \tag{5}$$

and $\mu$ is an integrable function satisfying

$$\mu \left( \sum_{i \in \{X,Y,Z\}} S_i u_i - V_0 \right) = 0, \quad \mu(t) \geq 0 \ a.e. \tag{6}$$

$$n_i(t) = -\lambda_i^S S_i - \mu S_i \ a.e. \text{ where } n_i(t) \in N_{[0,1]}(u_i^*(t)) \tag{7}$$

where $N_{[0,1]}$ denotes the normal cone to $[0, 1]$.

PMP then states that $u^* \in \text{argmax } \mathcal{H}$ where $u^*$ refers to an optimal control function. Note that the control variable is only included in the $\dot{S}_i$ term of the Hamiltonian. Thus, maximizing the Hamiltonian can be reduced to the following:

$$u^*(t) \in \text{argmax}_{u \in \mathcal{U}} \sum_{i \in \{X,Y,Z\}} -\lambda_i^S S_i u_i \tag{8}$$

where $\mathcal{U} = \{v : 0 \preceq v \preceq 1, \sum_{i \in \{X,Y,Z\}} S_i v_i \leq V_0\}$ denotes the set of admissible controls.
Let $a(t), b(t), c(t) \in \{X, Y, Z\}$ be distinct such that $-\lambda_{a(t)}^S S_{a(t)} \geq -\lambda_{b(t)}^S S_{b(t)} \geq -\lambda_{c(t)}^S S_{c(t)}$. Subject to choosing a control in $\mathcal{U}$, (8) implies that $u_{a(t)}^*(t)$ be maximized, subject to the same $u_{b(t)}^*(t)$ be maximized and so on. We now elaborate three cases.

1. Case 1: $S_{a(t)} \geq V_0$
   In this case, there are enough susceptible individuals in the highest priority group to fully consume vaccine capacity. Thus, $u_{a(t)}^*(t) = \frac{V_0}{S_{a(t)}}$ and $u_{b(t)}^*(t) = u_{c(t)}^*(t) = 0$.

2. Case 2: $S_{a(t)} < V_0 \leq S_{a(t)} + S_{b(t)}$
   Here the vaccine capacity exceeds the size of the susceptible population in the highest priority group and excess capacity is passed to the second priority group. Thus, $u^*_{a(t)}(t) = 1$, $u^*_{b(t)}(t) = \frac{V_0 - S_{a(t)}}{S_{b(t)}}$, and $u^*_{c(t)}(t) = 0$.

3. Case 3: $S_{a(t)} + S_{b(t)} < V_0$
   In this case, the vaccine capacity exceeds the total susceptible population of the first two priority groups, and any further vaccines are allocated to the final group until no further susceptible individuals remain. Thus, $u^*_{a(t)}(t) = u^*_{b(t)}(t) = 1$ and $u^*_{c(t)}(t) = \min\left(\frac{V_0 - S_{a(t)} - S_{b(t)}}{S_{c(t)}}, 1\right)$

This directly leads to Theorem 4.1 considering that $U_i(t) = u_i(t)S_i(t)$ for each $i, t$.

### 4 Numerical Evaluations

#### 4.1 Model Validation

We first describe how we obtain the parameters of our model for validation as described in Section 4.1. We consider publicly available data on infection and mortality counts in all US states and 139 countries [21]. Overall we considered a period extending from 04/01/2020 to 01/01/2021, the period between early reported deaths and the introduction of vaccines. We assume all initially infected individuals are in the baseline group. We obtain the initial values of the states from the publicly available infection counts on 04/01/2020 and the sizes of the different groups. We fix all individuals in age groups exceeding sixty as high risk. We obtain the fraction of the population in this group from publicly available population demographics [21]; this is the fraction of the population that is high risk. Percentage of workers characterized as essential workers[1] in US states is available in publicly available databases [23]. We use this fraction times the fraction of individuals of working age (20-60 years) as the fraction of the population that is high contact. Note that this estimate undercounts the fraction of high contact individuals as this does not consider unorganized private sector employees such as rideshare drivers who have high contact rates. For countries other than the US, we estimated the fraction of high contact individuals from economic data (percentage of GDP in service industries) and demographics (age-stratified population counts). That is, if $x\%$ of the country's economy was in the service industry (retail trade, transportation, and real estate being the largest share), then the high contact population was set to $x\%$ of people aged 20 to 60 [24]. We obtained disease parameters from CDC and WHO (Table 1, Supporting Information). After choosing the parameters as above, we determine the contact rate matrix using regression. The matrix consists of contact rates within each group (baseline, high risk, high contact) and across groups. These contact rates were selected to minimize mean squared normalized error (MMSNE) between the true infection and death counts in the respective area, and those projected for these locations via our model. We selected different sets of contact rates for different periods, each with a duration of two months (encompassing the time of investigation from 04/01/2020 to 01/01/2021). Different contact rates were chosen for different periods to account for changes in both government policy (e.g. start, relaxation, and end of lockdowns) and school openings which happen at low frequency.

**Model Validation Outliers** When regressing against actual infection and death counts [21], there were four countries (out of the total 139) that had outlying high MMSNE: Japan (JP), Lesotho (LS), Gambia (GM), and Mauritania (MR). While the max MMSNE of all other countries was 0.05, these four countries had MMSNE of 2.08, 10.16, 4.92, and 10.10, respectively. Here we seek to understand the factors that led to the poor fit. Each following subsection illustrates the fit of our model to the respective country's infection and death counts and posits feasible explanations for the high mismatch.

Japan is notable as the one country of the four outliers with a relatively high population size. In fact, Japan is the only of the four included in the top 125 countries by population [25]. As such, one would expect to see convergence between the system of ODEs and real data based on the Central Limit Theorem. However, the available case count data for Japan is flawed as the cumulative infection decreases over time which is impossible in practice. This artifact of the data collection did not allow us to fit real data. We therefore do not consider Japan.

Lesotho, with a population just over 2.2M, exhibits very low case counts and deaths [25]. As shown in Figure S3, this leads to non-smooth behavior in the real data which inhibits our ability to fit our model well.

Next, we consider Gambia. In addition to low overall counts, Gambia's data shows a sharp, sudden increase in death count as seen in Figure S4b. This is likely an artifact of delayed or imperfect record keeping of COVID-19 deaths and inhibits the fitting of smooth system dynamics.

[1] This group is composed of those working in critical industries who are not able to self isolate such as child care, energy, and transportation [22].

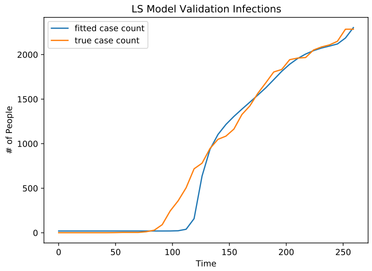


(a) Our model fit to Lesotho's infection counts

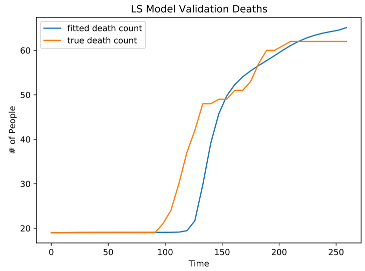


(b) Our model fit to Lesotho's death counts

Figure S3: Lesotho COVID-19 Dynamics Fitting

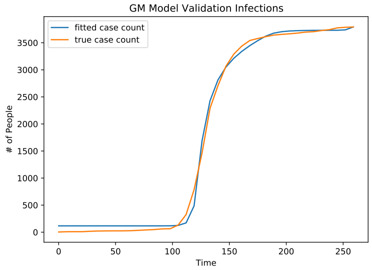


(a) Our model fit to Gambia's infection counts

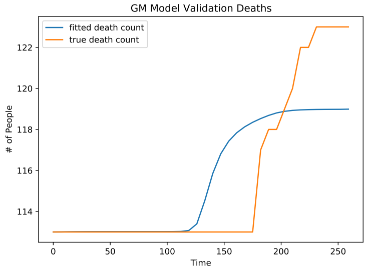


(b) Our model fit to Gambia's death counts

Figure S4: Gambia COVID-19 Dynamics Fitting

Mauritania, though it has a small number of cases, does exhibit relatively smooth case and death curves. However, at the onset of our time window, Mauritania had very few confirmed cases. Thus, with low initial infections, possibly due to undercounting, we see that our model shows delayed infection and death curves in Figure S5.

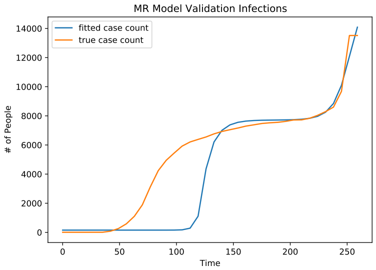


(a) Our model fit to Mauritania's infection counts

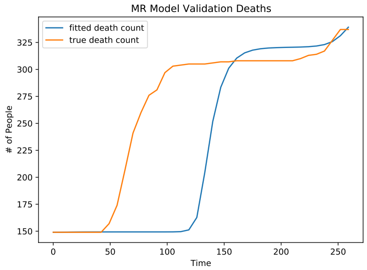


(b) Our model fit to Mauritania's death counts

Figure S5: Mauritania COVID-19 Dynamics Fitting

#### 4.2 Enumeration of Parameters Used for Numerical Evaluations

The sizes of different groups are determined as specified in Section 4.1. Contact rates between these groups were determined in one of two ways: (1) via modified survey-based contact matrices from [26] or (2) by regressing our model to fit real world infection and death counts from [21]. The latter case is described in Section 4.1. Here we discuss the former.

Country-specific contact matrices are obtained from [26]. These are provided in age increments of five years. To produce our contact matrices, we take the contact rate of the high risk group to be the weighted average of the contact rates of each 65+ age group with weight corresponding to the population of each age group obtained from [21]. The contact rate of the high contact group is set to be a multiple of that of the average contact rate over working age individuals. The contact multiple for the high contact group is varied between 1.2 and 6. Finally, $R_0$ is computed via the next generation matrix method and normalized (by scalar multiplication of the contact matrix) to a fixed value in our parameter range (following the methodology of [27]). By default we set the vaccination capacity constraint to be at 0.5% of the total population daily unless otherwise specified [28].

We obtain the demographic data from census data as in the previous section. We consider the level of initial infection specified as the fraction of the overall populace. Initial infections are varied between 0.1% and 1% – this range captures the seroprevalence upon the introduction of vaccines in all US States and nearly all countries for which data was available[2] [21]. The default assumption is that initial infections are seeded only in the baseline group; we explicitly specify when we deviate from the default assumptions. We fix the efficacy of vaccine at preventing infection, symptoms, hospitalization, and death as 50%, 70%, 80%, and 90%, respectively unless otherwise specified [12]. We used the contact rates obtained from the real evolution of the pandemic as in the previous section. We also use additional contact rates from ranges of contact rates within and across age groups in 139 countries obtained from surveys and the sizes of the different groups [26]. Over all, the large range of contact rates we consider capture varying degree of implementation and compliance with non-pharmaceutical interventions (NPIs) such as social distancing and lockdowns in different US states and the 139 countries. Finally we vary the vaccine efficacy, that is, the rates at which vaccinated individuals become infected, symptomatic, hospitalized, and deceased, over ranges drawn from clinical studies of various vaccines [12, 13]. Over all these parameter ranges, our model was instantiated and run on a fine grid of approximately 911,250 settings. The associated ranges are included below and are used for the results detailed throughout Sections 4.2, 4.3, 4.5, and 4.6.

Table 4: Model Instances

| Parameter | Setting(s) | Ref. |
|---|---|---|
| Contact matrices | 150 country estimates | [26] |
| COVID-19 Variants | {alpha, delta, omicron} | [30], [31] |
| $R_0$ | {1, 1.5, 2, 2.5, 3} | [32] |
| NPI Efficacy | {0, 0.3, 0.6} | [33] |
| Initial Infections | [0.1%, 0.25%, 0.5%, 0.75% 1%] | [21] |
| Vaccine Efficacy | baseline, baseline ± 20% | [12],[13] |
| High Contact Population Size | {10%, 15%, 20%} | [23] |
| Transmisibility Multiplier | {1, 1.5, 2} | [34] |

For our case studies in Section 4.4, we depart slightly from the above methodology where necessary. In our case study of LMICs, the vaccination capacity constraint is tightened to 0.2% to reflect scarcity [28]. For the prison and nursing home case studies, population demography was obtained from the Bureau of Prisons and the CDC, respectively [35, 36]. Contact matrices for the US Baseline and LMIC were obtained as specified above. For prisons and nursing homes, contact rates were obtained from [37] and [38, 39], respectively.

[2]There is a single country (Gibraltar) which had an initial infection rate higher than our range. In addition, there are countries which are relatively isolated and/or have low population (i.e. New Zealand) that have initial infection rates below our range. Finally, there are a few large countries, notably India and China, which fall below this range, but this may be attributed to suspected undercounting of cases [29].

#### 4.3 Robustness Study

The estimates of parameters, particularly the disease parameters, will inevitably have some errors. We therefore investigate the robustness of the computation framework for the optimal vaccination strategy to estimation errors as mentioned in Section 4.2. Toward this end, we inject additive white Gaussian noise into transmissibility and fractions of individuals who become symptomatics (from the exposed state), are hospitalized (from late stage infection) and die (after hospitalization). That is, for each such parameter, the value was multiplied by $(1 + \mathcal{N}(0, \sigma^2))$ where $\mathcal{N}(\mu, \sigma^2)$ is the normal distribution with mean $\mu$ and standard deviation $\sigma$ [3] These parameters affect transition rates into some states. We took $\sigma^2 \in \{0.05, 0.1, 0.15\}$. While the true dynamics of the evolution were determined by the original parameter, the computation framework only had access to these noisy estimates. The optimal vaccination strategy computed using these noisy estimates is referred to as 'noisy optimal." The average increase in death count due to utilization of the noisy optimal strategy was below 2%, although, in extreme cases it was as high as 84.6%. Between high contact and high risk policies, the policy that has lower death counts was identified correctly under the noisy parameters in 89.1%, 88.2%, and 86.2% of cases, respectively for $\sigma^2 \in \{0.05, 0.1, 0.15\}$. Thus, overall the framework is robust to estimation error.

When evaluating the vaccination policy made with noisy information, 88.2% of all instances exhibited a subpotimality of less than 1% when the controller only had access to noisy disease parameters (noise powers of 5%, 10%, and 15%). The histogram of the suboptimality in the remaining 11.8% of cases is included below in Figure S6.

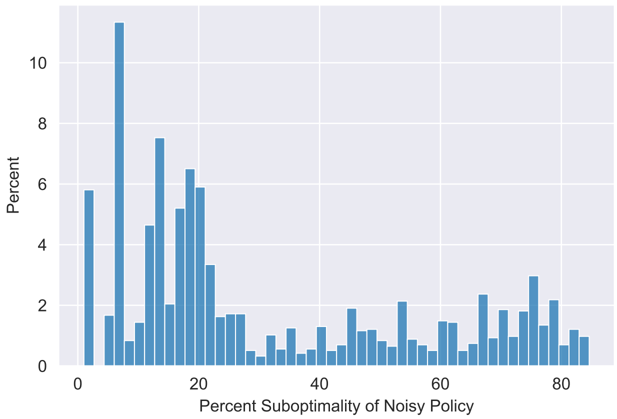


Figure S6: Histogram of suboptimality of noisy vaccination policy when suboptimality exceeds 1%

[3] We ignore the few cases in which the erroneous estimates become negative. Whenever the erroneous estimate of the fractions exceed 1, we consider the fraction to be 1.

### 5 Runtime Performance and Comparison

Throughout our paper we refer to the computational efficiency of our framework (see Sections 2, 4.2, 5). Here we evaluate the runtime of computing the optimal vaccination policy over the landscape of instances defined in Section 4.2. As shown in Figure S7, the runtimes of our algorithm, omitting the small number of instance that reached the cutoff time of 500 seconds, were heavily concentrated under 10 seconds. This is a drastic improvement over simulation based techniques or more complex optimization protocols and allows us to survey broad parameter landscapes to map optimal policies as presented in Section 4.3.

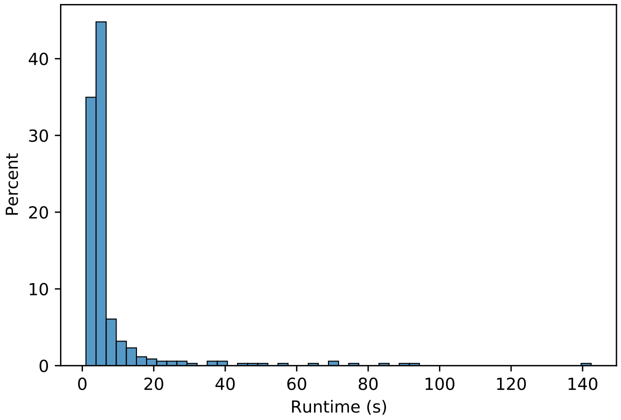


Figure S7: Histogram of runtimes for our optimal control-based vaccine prioritization method

To our knowledge, the closest work to consider the optimal selection of dynamic COVID-19 vaccination policies is the paper authored by Buckner et. al. [40]. Here they utilize a genetic algorithm based on the work in [41] followed by a simulated annealing algorithm to more precisely identify their optimal. Genetic algorithms and simulated annealing can optimize only discrete variables but not arbitrary functions of time. Thus these can optimize vaccination strategies only among a reduced strategy space, namely, among strategies that allocate piece-wise constant vaccination rates to different groups. Specifically, the overall time horizon under consideration is partitioned into intervals and the fraction of overall capacity allocated to different groups in each interval are considered as constants over time in the interval. The fractions in different intervals are the optimization variables, which are chosen optimally, to optimize the public health objective in this restricted policy space. The policy space becomes closer to the space of all vaccination strategies as the number of intervals increase. But the computation time for the genetic algorithm is exponential in the number of time intervals [41]. Thus the computation time increases rapidly as the number of intervals increase. Buckner et. al. considered a time horizon of 6 months and the interval size of 1 month; thus the vaccination rates across the groups they consider can change only once a month [40]. But even with such coarse granularity their genetic algorithm step alone can take hours to converge to an optimal in a single instance, even without including the subsequent simulated annealing. Since the computation time is exponential in the number of time intervals, runtime becomes intractable when considering highly dynamic policies as shown in Table 5, which rules out obtaining solutions using current computing capabilities and thereby using such solutions even as benchmarks for comparison let alone for actual deployment. This is an inherent limitation on the ability of genetic algorithms to consider highly dynamic policies. In contrast, our computation framework optimizes the vaccination policy in much broader policy space than the above, among policies whose choices can arbitrarily vary at any time granularity; still our framework yields the optimal policy in seconds. Even considering policies that can change allocations only once every month, Buckner et. al. presented the optimal in this reduced space only for a small number of instances [40],

| Runtime Comparison | | |
|---|---|---|
| Length of Decision Intervals (days) | Our Method Mean Runtime (s) | Buckner et al. Mean Runtime (s) |
| 90 | 2.86 | 2470.71 |
| 30 | 9.59 | 3076.86 |
| 10 | 10.50 | 6277.54 (projected) |
| 7 | 14.54 | 8258.43 (projected) |
| 1 | 160.59 | $4.30 \times 10^7$ (projected) |

Table 5: This table shows the mean runtime of determining the optimal vaccination policy when instantiated on different contact matrices and demography [21, 26]. For shorter decision interval lengths, the runtime of Buckner et al.'s algorithm is projected using an exponential fit with the number of decision variables as the independent variable because their algorithm is exponential in this dimension.

perhaps due to the long computation time of their method, whereas we could present results for large landscapes of 911,250 instances involving variations of parameters in large realistic ranges (Section 4). Large landscapes allow for more reliable conclusions on how the optimal strategy and the optimal value of the public health objective changes with variations in parameter values; large landscapes also help identify near-optimal easy-to-deploy vaccination strategies which we accomplish. Next, generalizations to consider several attributes of practicality inevitably increases the state space of the set of trajectory (e.g. when breakthrough infections and reinfections are considered), and in many instances the number of functions that need to be optimized (e.g. for two dose vaccination). All these increase the computation time for each instance further and therefore renders the computations even more formidable if they are involved in the simplest case. Thus computational tractability of the basic framework is imperative for its adaptability to more involved albeit realistic settings. Next, most works in this genre (including Buckner *et al* partition the populace into a larger number of groups than us, and based on a different criteria (age and in one case profession) [40, 27]. Our computation time for each instance is lower also because we consider fewer groups (though the primary reason is the choice of the optimal control formulation rather than the genetic algorithm and simulated annealing combo). As mentioned before, our model is able to accurately predict the evolution and spread of COVID-19 in geographical units of different scale and locations all over the world, despite consideration of partition criteria that yields fewer groups. Finally, our optimal strategy is structurally different from that obtained by Buckner *et al* because of 1) the significant difference in time granularity over which vaccination rates allocated to different groups can change (ours can change any time, theirs can change only once a month) and 2) different criteria for partition of the populace into groups. For example, the policy obtained by Buckner et al is always mixed, i.e., vaccinates multiple groups simultaneously, whereas we analytically argue that the death count does not increase if the policy space is limited to those that vaccinate one group at a time, while the susceptibles in each group exceeds the vaccine capacity (i.e. while each group has enough individuals to vaccinate if it is given the full vaccination capacity). If we optimize in a restricted policy space where the allocations can change only once a month, our optimal strategy is also mixed from the start.

#### References


[1] *Coronavirus disease (covid-19): How is it transmitted?* URL: https://www.who.int/news-room/questions-and-answers/item/coronavirus-disease-covid-19-how-is-it-transmitted.

[7] *Interim clinical guidance for management of patients with confirmed coronavirus disease (covid-19).* URL: https://stacks.cdc.gov/view/cdc/88624.

[8] *Mortality analyses.* URL: https://coronavirus.jhu.edu/data/mortality.

[9] *CDC COVID-19 study shows mrna vaccines reduce risk of infection by 91 percent for fully vaccinated people.* June 2021. URL: https://www.cdc.gov/media/releases/2021/p0607-mrna-reduce-risks.html.

[10] National Center for Immunization and Respiratory Diseases. "Science Brief: SARS-CoV-2 Infection-induced and Vaccine-induced Immunity". In: *CDC COVID-19 Science Briefs [Internet]*. Centers for Disease Control and Prevention (US), 2021.

[11] *Comparing the differences between covid-19 vaccines.* URL: https://www.mayoclinic.org/coronavirus-covid-19/vaccine/comparing-vaccines.

[19] Lev Semenovich Pontryagin. *Mathematical theory of optimal processes.* CRC press, 1987.

[20] Dieter Grass et al. *Optimal control of nonlinear processes with applications in drugs,corruption, and terror.* Springer, 2010.

[21] O. Wahltinez et al. "COVID-19 Open-Data: curating a fine-grained, global-scale data repository for SARS-CoV-2". In: (2020). Work in progress. URL: https://goo.gle/covid-19-open-data.

[22] *Report COVID-19: Essential Workers in the States.* URL: https://www.ncsl.org/labor-and-employment/covid-19-essential-workers-in-the-states.

[23] *US states with the most essential workers.* Dec. 2021. URL: https://unitedwaynca.org/blog/us-states-with-the-most-ssential-workers/.

[24] URL: https://bbs.portal.gov.bd/sites/default/files/files/bbs.portal.gov.bd/page/057b0f3b_a9e8_4fde_b3a6_6daec3853586/2021-12-02-10-01-a5b3adcd2ea20db89d4bae0c90bd86cf.pdf.

[25] *Population, total.* URL: https://data.worldbank.org/indicator/SP.POP.TOTL.

[26] Kiesha Prem, Alex R Cook, and Mark Jit. "Projecting social contact matrices in 152 countries using contact surveys and demographic data". In: *PLoS computational biology* 13.9 (2017), e1005697.

[27] Kate M Bubar et al. "Model-informed COVID-19 vaccine prioritization strategies by age and serostatus". In: *Science* 371.6532 (2021), pp. 916–921.

[28] Edouard Mathieu et al. "A global database of COVID-19 vaccinations". In: *Nature human behaviour* 5.7 (2021), pp. 947–953.

[29] Claire Klobucista. *By how much are countries underreporting COVID-19 cases and deaths?* URL: https://www.cfr.org/in-brief/how-much-are-countries-underreporting-covid-19-cases-and-deaths.

[35] *Federal Bureau of Prisons.* URL: https://www.bop.gov/about/statistics/population_statistics.jsp.

[36] *FASTSTATS - Residential Care Community.* Dec. 2022. URL: https://www.cdc.gov/nchs/fastats/residential-care-communities.htm.

[37] Martial L Ndeffo-Mbah et al. "Dynamic models of infectious disease transmission in prisons and the general population". In: *Epidemiologic reviews* 40.1 (2018), pp. 40–57.

[38] Andrew T Levin et al. "COVID-19 prevalence and mortality in longer-term care facilities". In: *European Journal of Epidemiology* (2022), pp. 1–8.

[39] Courtney H Van Houtven, Nathan A Boucher, and Walter D Dawson. "Impact of the COVID-19 outbreak on long-term care in the United States". In: *International Long-Term Care Policy Network* (2020).

[40] Jack H Buckner, Gerardo Chowell, and Michael R Springborn. “Dynamic prioritization of COVID-19 vaccines when social distancing is limited for essential workers”. In: *Proceedings of the National Academy of Sciences* 118.16 (2021).

[41] Rajan Patel, Ira M Longini Jr, and M Elizabeth Halloran. “Finding optimal vaccination strategies for pandemic influenza using genetic algorithms”. In: *Journal of theoretical biology* 234.2 (2005), pp. 201–212.
